# Remote Assessment of Cognition in Kids and Adolescents with Daytime Sleepiness: A pilot study of feasibility and reliability

**DOI:** 10.1101/2021.03.24.21254190

**Authors:** Jennifer Worhach, Madeline Boduch, Bo Zhang, Kiran Maski

## Abstract

In this pilot study, we assessed the reliability of cognitive testing for kids and adolescents ages 8-19 years of age with narcolepsy or subjective daytime sleepiness compared to healthy controls. Forty-six participants took part in the study (n=18 with narcolepsy type 1, n=6 with subjective daytime sleepiness, and n= 22 healthy controls recruited from the community). Participants completed verbal (vocabulary testing) and non-verbal intelligence quotient (IQ) tasks (block design, matrix reasoning) from the Weschler Abbreviated Scale of Intelligence-Second Edition (WASI-II) in-person or remotely in their home through a HIPAA compliant telehealth web platform with conditions counterbalanced. We found that vocabulary T-scores showed good reliability with intraclass correlation coefficient (ICC) of 0.76 (95% CI: 0.64, 0.85) between remote and in-person testing conditions. Matrix Reasoning T-scores showed moderate reliability (ICC 0.69, 95% CI: 0.68, 0.90) and Block Design T-scores was poor between testing conditions. Bland-Altman plots showed outliers on vocabulary and matrix reasoning tasks performed better on remote assessments. Overall, the results of this pilot study support the feasibility and reliability of verbal and non-verbal IQ scores collected by telehealth. Use of telehealth to collect verbal and non-verbal IQ scores may offer a means to acquire cognitive data for pediatric sleep research through the COVID-19 pandemic and beyond.

## Introduction

Telehealth has expanded exponentially in many areas of healthcare over the past decade and continues to grow in the setting of the COVID-19 pandemic. Telehealth also holds great promise for research, especially in the assessment of cognitive measures. Measure of objective intellectual function are of great interest in sleep medicine as cognitive difficulties are frequently reported people with sleep disorders such as narcolepsy and obstructive sleep apnea (Blackwell et al., 2017; Naumann et al., 2006; Stranks & Crowe, 2016; Maski et al., 2017; Vaessen et al., 2015). Given the challenges of assessing cognition in-person due to access to transportation, time burden for patients and caregivers, facility limitations, and current concerns of viral transmission, validating remote cognitive assessments participants with sleep disorders is of great value to facilitate future research endeavors.

Reliability of remote cognitive testing has been best studied in adult patients and has included healthy individuals as well as those with mild cognitive impairment, Alzheimer’s disease, a history of alcohol abuse, and intellectual disabilities (Cullum et al., 2014; Cullum et al., 2006; Jacobsen et al., 2003; Kirkwood et al., 2000; Temple et al., 2010). Results have shown that remotes assessments yield comparable results to face-to-face assessments. In general, studies have also found participants are accepting and comfortable with the use of technology in performing cognitive assessments (Hildebrand et al., 2004; Parikh et al., 2013). Fewer pediatric studies exist assessing reliability of remote cognitive testing but include assessments of children with Batten disease, learning disabilities, and psychosis (Hodge et al., 2019; Ragbeer et al., 2016; Stain et al., 2011). However, none of the remote testing sites in these pediatric studies were conducted in home settings but rather hotel rooms during disease based conferences or within a designated testing site with patients in one room and assessor in another. Theoretically, remote cognitive testing in the home environment could pose more challenges as kids and adolescents may be more distracted in home settings.

In this pilot study, we assess the reliability of a subset of verbal and non-verbal IQ tests from the Weschler Abbreviated Scale of Intelligence-Second Edition (WASI-II) done via home-based telehealth services against in-person testing among healthy controls and participants with subjective sleepiness (patients presenting with subjective excessive daytime sleepiness but had normal polysomnogram and multiple sleep latency testing) or narcolepsy type 1 (NT1) aged 8-19 years. In addition, we examined the influence of self-reported subjective sleepiness and sleep, affect, circadian preference, and objective measures of sleep duration from actigraphy on cognitive test results. We hypothesized that remote cognitive assessments would show good reliability with in-person testing across all groups and task scores collected in the different testing conditions would not show associations with sleepiness, sleep, nor affect measures.

## Methods

### Participants

We recruited participants from the Boston Children’s Hospital sleep clinic through clinic flyers and web ads and the community through the Boston Children’s Hospital Research Patient Registry, a recruitment database of potential subjects who have indicated that they are interested in hearing about research studies. In total 46 participants ages 8 to 19 took part in the study (see Table 1 for demographic and medication history). Participants included 18 children with Narcolepsy Type 1 (NT1) (American Academy of Sleep Medicine, 2014), 6 with subjective sleepiness (patients who reported symptoms of excessive daytime sleepiness but did not meet clinical criteria for a CNS Disorder of Hypersomnolence based on polysomnogram and multiple sleep latency testing) (American Academy of Sleep Medicine, 2014), and 22 healthy controls recruited from the community. During remote testing, NT1 participants were allowed to take their home medications as prescribed, but for in-person testing, participants were asked to stop stimulants, wake promoting medications, or any sedating medications for 5 half-lives of the drug. NT1 participants were permitted to stay on SSRI medications for cataplexy.

**Table 1:** Demographics of Participants.

| Characteristics | Healthy Controls<br>N=22 | Subjectively Sleepy<br>N=6 | Narcolepsy Type 1<br>N=18 | p-value |
| --- | --- | --- | --- | --- |
| Age | 13.59 (3.1) | 14.00 (3) | 14.94 (2.4) | .32 |
| BMI | 23 (4.8) | 20.6 (4.3) | 28.9 (6.6) | .001 |
| Female | 10 (52.6) | 1 (5.3) | 8 (42.1) | .42 |
| Male | 12 (44.4) | 5 (18.5) | 10 (37.0) |  |
| Caucasian | 15 (68.2) | 4 (66.7) | 9 (50) | .43 |
| African American | 2 (9.1) | 1 (16.7) | 6 (33.3) |  |
| Asian | 1 (4.5) | 1 (16.7) | 1 (5.6) |  |
| Other Ethnicity | 4 (18.2) | 0 (0.0) | 2 (11.1) |  |
| Maternal Education<br>High School | 3 (13.6) | 0 (0.0) | 3 (16.7) | .76 |
| Maternal Education College | 9 (40.9) | 4 (66.7) | 8 (44.4) |  |
| Maternal Education<br>Grad School | 10 (45.5) | 2 (33.3) | 7 (38.9) |  |
| Stimulant/wake promoting<br>medication withdrawal | 0 (0.0) | 2 (33.3) | 12 (66.7) | .000 |
| On SSRI/SNRI | 0 (0.0) | 0 (0.0) | 7 (38.9) | .002 |

\*Mean and standard deviation (in brackets) are displayed for Age and BMI. Total number of participants and percentage (in brackets) are displayed for the other characteristics.

Written consent and assent was obtained from all participants prior to beginning the study.

### Protocol

The IQ measures collected in this pilot study were part of a larger clinical research study conducted at Boston Children’s Hospital. In order to assess baseline IQ, three trained research assistants independently administered the WASI-II subtests to participants once remotely and once in person with conditions counterbalanced to negate practice effects. We used WASI-II vocabulary testing task to assess verbal IQ and either block design or matrix reasoning task for non-verbal IQ assessment. Remote testing occurred via HIPAA compliant platforms, Zoom Video Communications, Inc. and Vidyo, Inc., with the assessor in their office or home and participant in their home. Participants were mailed a sealed packet of testing materials for WASI-II tasks for the remote assessment and broke the seal of the packet materials in front of the examiner during the remote testing session on web-camera. Both Vidyo, Inc. and Zoom Video Communications, Inc. provide a secure communication environment with Advanced Encryption Standard (AES) and are compliant with the data security requirements of the Health Insurance Portability Act and Accountability Act. Remote testing occurred in the late afternoon (between 3-4:30 pm) for all participants (after school) except for two participants whose testing occurred in the morning (9:30am - 10:00am) due to scheduling conflicts. In-person testing occurred at Boston Children’s Hospital in a designated testing room between 6 pm-7 pm one week before or after the remote assessment (conditions counterbalanced, chi-square value 2.54, p=0.28). For each participant, the same research assistant conducted both testing conditions using a personal laptop computer. Research assistants followed standard administration procedures across conditions which included affirming both parties web cameras were working with good picture clarity, no distractions present, and comfort at time of testing. Testing proceeded after participants verbally affirmed these conditions. The remote and in-person cognitive assessments took between 25-45 minutes for participants to complete in either condition (home/remote).

Prior to in-person cognitive testing, we sent participants 4 questionnaires regarding affect, chronotype, sleep problems and daytime sleepiness via REDCap (Harris et al., 2009) survey distribution system and they wore an actigraph (Actiwatch-L, Mini-Mitter/Respironics) on their non-dominant wrist for 7 days to assess habitual total sleep time.

Institutional Review Board approval was obtained from the Boston Children’s Hospital.

### Measures

Weschler Abbreviated Scale of Intelligence Second Edition (WASI-II): To assess participants baseline verbal and non-verbal IQ T-scores, we used the WASI-II. This is a widely used measure of objective cognitive ability that consists of four subtests (Wechsler, 2011). To assess baseline verbal IQ, we administered the vocabulary task for all participants and to assess non-verbal IQ, we administered either the block design or matrix reasoning subtests. We used T-score generated from this testing to assess reliability measures. Subtest T-scores mean is 50 and standard deviation is 10 (normative range).

Positive and Negative Affect Scale for Children (PANAS-C): This is a 30 item self-report questionnaire with 15 negative affect items and 12 positive affect items. The PANAS–C instructs children to indicate how often they have felt interested, sad, etc. during the “past few weeks” on a 5-point Likert scale. It is validated in children ages 10-18 years (Laurent et al., 1999; Watson et al., 1988).

Children’s Sleep Habits Questionnaire (CSHQ): This is a 45 item questionnaire filled out by the parent to screen for underlying sleep problems. Parents are asked to rate the frequency in which their child displayed various sleep behaviors over the past week. It is validated in children ages 4 -10 years (Owens et al., 2000).

Morningness and Eveningness Scale for Children (MESC): This is a 10 item self-reported questionnaire used to evaluate morning/evening sleepiness preferences in children that may influence cognitive testing. It examines sleep schedule inclinations and subjective feelings of fatigue and alertness. It is validated in children as young as 9 years of age (Carskadon et al., 1993).

Adapted Epworth sleepiness scale (ESS): This is an 8 item self-reported questionnaire used to assess for daytime sleepiness. It asks subjects to rate the probability of falling asleep during daytime situations and activities. It is validated in children 6-16 years (Janssen et al., 2017; Johns, 1991).

Actigraphy testing: Actigraphy is a wrist-watch device worn on the non-dominent wrisr that has a computer-based validated algorithm to measure wake and sleep periods in children and adults based on movement (Morgenthaler et al., 2007). It is currently the gold standard for home ambulatory sleep/wake measurement and validated in children (Meltzer et al., 2016). We report one week average total sleep time (TST) measures collected from the (Actiwatch-L, Mini-Mitter/Respironics device).

### Statistical Analysis

To compare demographic variables, survey data and actigraphy results, we used ANOVA testing. To report WASI-II task scores, we report mean scores in each condition (remote, in-person) for each group and conducted a univariate screen with ANOVA testing for group differences. If ANOVA testing yielded results with p-value <0.01, we conducted linear regression for group differences adjusting for age, gender and ethnicity.

WASI –II task scores showed normal distribution. We used paired t-tests to compare the mean difference between scores collected in the two testing conditions.

To look at the relationship between WASI-II subtest scores and sleepiness, affect and sleep variables, we conducted Pearson correlation testing with generated T-scores with the following variables: CSHQ total score, morning/eveningness scores, ESS total score, TST average on actigraphy, and positive and negative mood scales. If these correlations showed p-value of <0.01, we then further analyzed that association in a linear regression model adjusting for age, gender, and ethnicity.

We assessed reliability between the two methods of administration (telehealth vs. in person) both within and across groups using intraclass correlations coefficients (ICC). ICC is an index that assess not only how well correlated two techniques are but also if they are equal. ICC ranges from 0 (no agreement) to 1 (perfect agreement). An ICC <0.5 indicates poor agreement, 0.5-0.75 indicates moderate agreement and 0.75 -0.9 indicates good agreement and >0.90 indicates excellent agreement (Koo & Li, 2016). We used Bland-Altman plots to assess the mean differences between measures and visually examine the degree of agreement between the two testing conditions. In this method, the differences between remote and in-person testing are plotted against their averages. The plot includes one value per participant, a reference line (zero representing perfect agreement between testing conditions), the mean of the differences between the conditions (mean bias), and limits of agreement (deviation from the mean superior to two standard deviations). Differences are expressed as remote assessment minus in-person assessment so a negative value indicates that in-person has a higher IQ score than the remote assessment and a positive value indicates remote assessment overestimates the value. Bland and Altman recommended that 95% of the data points should lie within ± two standard deviations of the mean difference to assess reliability{Bland, 1999 #169}.

We conducted statistical analysis using SPSS for Windows (version 19; IBM Corp, Armonk, NY, USA).

## Results

Of the 46 participants, all completed the WASI-II vocabulary subtest. Only the first 12 participants completed the block design subtest because we found poor reliability on interim analysis and thus switched to the matrix seasoning subtest (n=34 participants) for the remainder of the study. Mean age of participants (14.21 years, SD 2.7) was similar between groups (F=1.26, p=0.29). Gender (chi-square 0.78, p=0.68) and ethnicity (chi-square 5.69, p=0.49) did not differ between groups either. Full demographic results are listed in **Table 1**. Task performance data, survey and actigraphy data results are listed in **Table 2**.

**Table 2:** Data from Cognitive Performance Tasks, Survey and Actigraphy.

| Measure | N Value | Healthy Controls Mean (SD) | N Value | Subjectively Sleepy Mean (SD) | N Value | Narcolepsy Type 1 Mean (SD) | P Value |
| --- | --- | --- | --- | --- | --- | --- | --- |
| CSHQ | 21 | 41.33 (5.58) | 6 | 50.83 (3.19) | 17 | 55.18 (5.25) | 0.00 |
| Remote Vocabulary T-score | 22 | 57.59(8.34) | 6 | 56.50 (7.23) | 18 | 52.94 (9) | 0.23 |
| Remote Matrix Reasoning T-score | 16 | 55.31 (11) | 2 | 41.50 (4.955) | 16 | 54.13 (7.74) | 0.16 |
| Remote Block Design T-score | 6 | 50.33 (6.06) | 4 | 51.25 (6.95) | 2 | 45 (2.83) | 0.50 |
| In-person Vocabulary T-score | 22 | 56.86 (6.40) | 6 | 55.33 (8.12) | 18 | 51.44 (6.24) | 0.04 |
| In-person Matrix Reasoning T-score | 16 | 56.25 (10.096) | 2 | 41.00 (1.41) | 16 | 54.44 (8.21) | 0.10 |
| In-person Block Design T-score | 6 | 53.83 (5.78) | 4 | 51.50 (7.04) | 2 | 52.50 (7.78) | 0.86 |
| ESS | 22 | 4.50 (3.71) | 6 | 10.83 (5.49) | 18 | 15.00 (2.6) | 0.00 |
| MESC | 22 | 29.91 (2.98) | 6 | 26.17 (6.71) | 18 | 26.78 (5.91) | 0.08 |
| PANAS-C (Negative scale) | 22 | 20.64 (4.95) | 6 | 26.83 (8.75) | 18 | 27.11 (10.98) | 0.04 |
| TST Actigraphy | 16 | 514.3 (48) | 4 | 533.8 (43.9) | 17 | 565.51(89) | 0.13 |

NOTE: Vocabulary, block design and matrix reasoning scores are from the Weschler Abbreviated Scale of Intelligence- Second Edition. CSHQ= Children's Sleep Habits Questionnaire; ESS = Epworth Sleepiness Scale; MESC= Morningness and Eveningness Scale for Children; PANAs-C = Positive and Negative Affect Scale for Children; TST= Total Sleep Time

### WASI-II task performances were similar between groups

All groups’ WASI-II task scores fell within normal range (normal range defined as mean T-score, SD 10). On ANOVA univariate screen, we detected group differences in in-person vocabulary task results but group differences did not persist when adjusting for age, gender and ethnicity (F=2.12, p=0.13).

### Remote testing of Verbal IQ shows good reliability with in-person testing

We assessed the reliability of the vocabulary T-scores using ICC and Bland-Altman plots and analysis (LoA). In the two-way mixed effects model of all participants, we found an ICC of 0.76 (95% CI: 0.64, 0.85) between remote and in-person administered vocabulary T-scores indicating good reliability. On subgroup analysis, the reliability for the subjectively sleepy controls (ICC 0.93, 95^%^ CI: 0.74, 0.98) was excellent and NT1 participants (ICC 0.82, 95^%^ CI: 0.63, 0.91) was good, but the reliability in the healthy control was only moderate (ICC 0.63, 95% CI: 0.35, 0.80). The Bland-Altman plot of vocabulary T-scores is presented in Figure 1a for all participants and each group (subjectively sleepy, narcolepsy type 1 and healthy controls). We identified three outliers (2 healthy controls and 1 NT1 participant) who had higher scores >2 SD on the remote assessment compared to in-person testing. As the healthy control group did show lower reliability, we reviewed data on the two healthy control outliers and did not find they differed from the group in demographics, actigraphy result, or survey responses.

**Figure 1.**
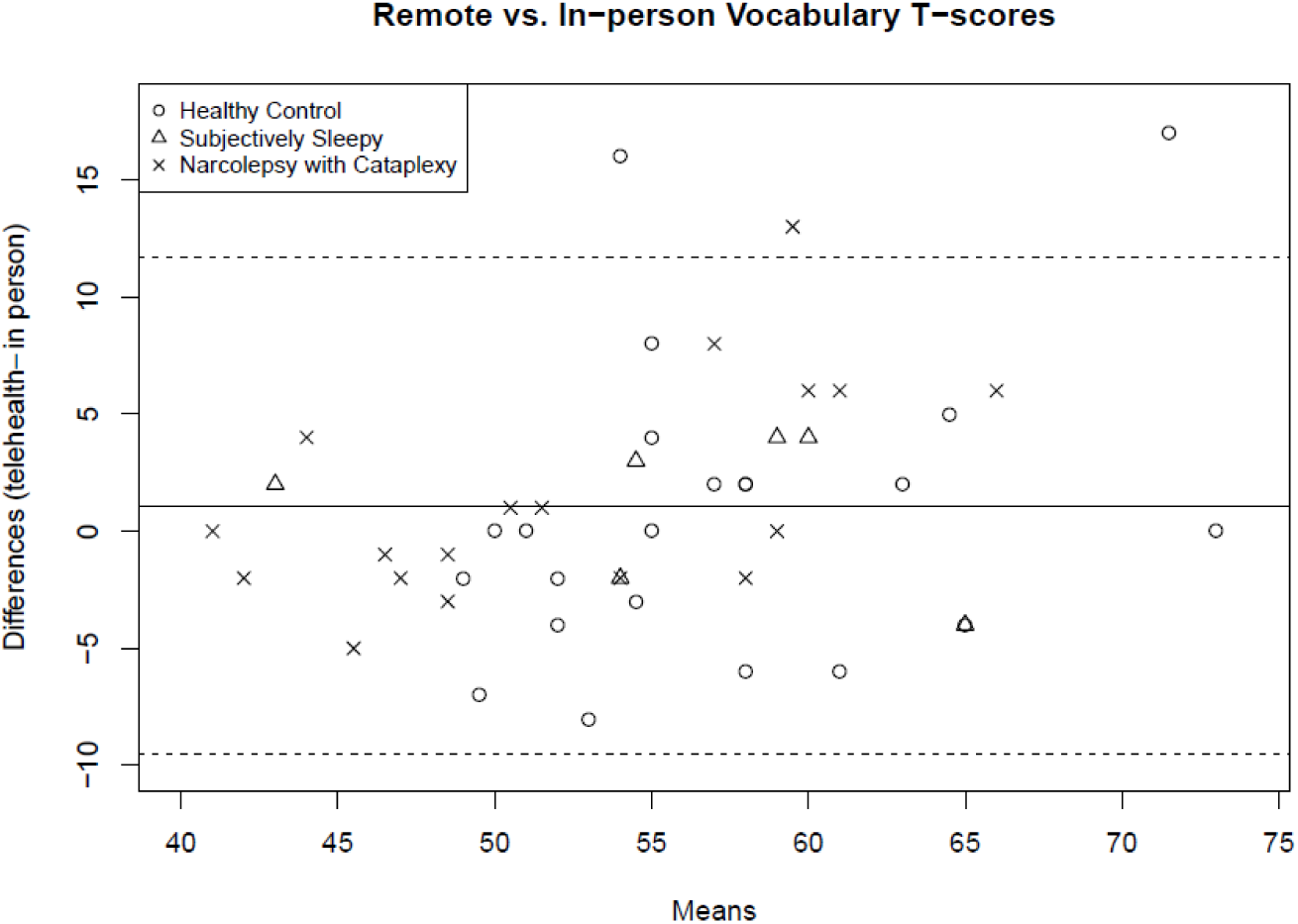
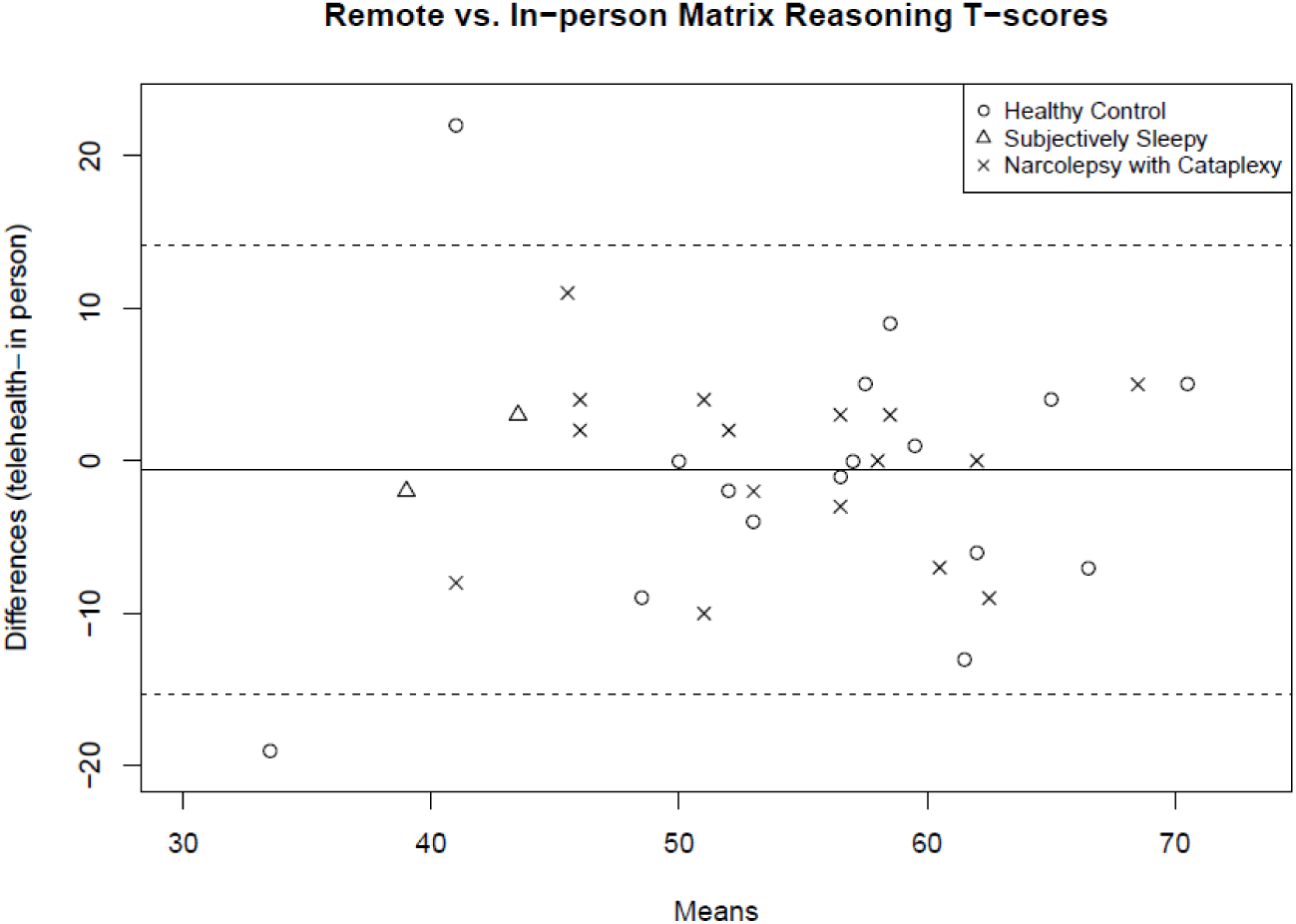
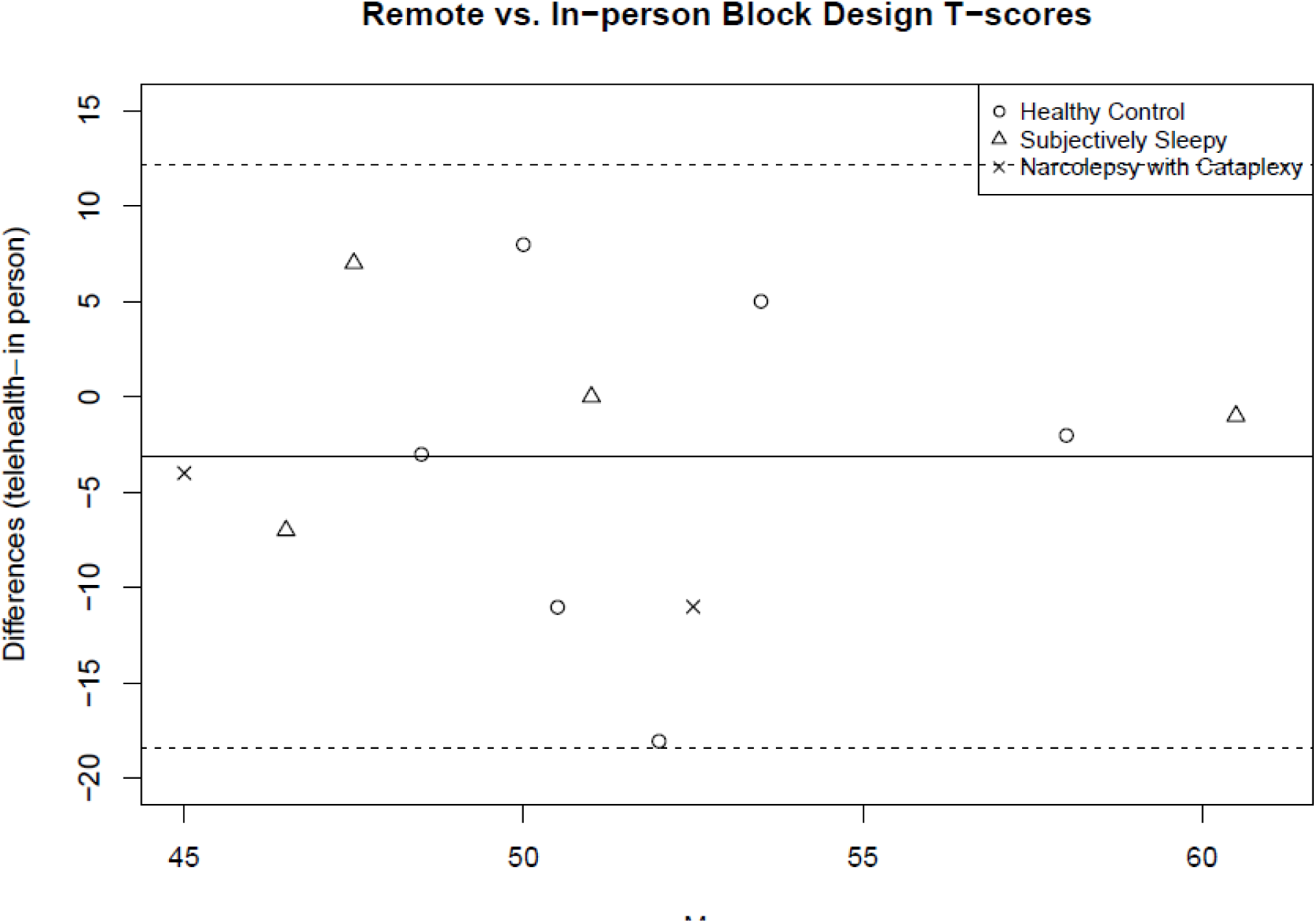
Bland Altman Plots for WASI-II Subtests Remote vs. In-person Conditions.

Across all groups, the in-person vocabulary T-scores was slightly less than those obtained by remote assessment but results were not significant (mean difference 1.1, p=0.18) based on paired t-test.

### Remote testing of WASI-II matrix reasoning task shows moderate reliability with in-person testing

Among the 34 participants who completed the in-person and remote matrix reasoning task from the WASI-II, our two-way mixed effects model showed moderate reliability of the T-scores between conditions [ICC 0.69, 95% CI: 0.68, 0.90). However, we found excellent reliability among the subjectively sleepy participants with ICC of 0.93, 95% CI: 0.71,0.98) whereas the other groups showed only moderate reliability (NT1: ICC 0.55, 95% CI: 0.32, 0.72; healthy controls: ICC 0.71, 95% CI:0.53,0.82). The Bland-Altman plots are presented in Figure 1b for all participants and show that two healthy controls were outliers. One of these outliers performed better on remote testing condition and the other performed better in-person.

Based on paired t-tests there was no significant difference across participants on their in-person vs. remote matrix reasoning T-scores (mean difference -0.56, SD 7.51, p=0.67).

### Remote testing of block design shows poor reliability with in-person testing

Only 12 participants completed the block test because reliability was clearly poor between testing conditions during this pilot study. Across groups, the ICC was 0.14, 95% CI 0, 0.56 and there were too few participants in each group to perform meaning subgroup analysis. The Bland-Altman plots are presented in Figure 1c for all participants and show that all participants had results within two standard deviations of the mean. Paired t-tests did not show differences between conditions (mean difference -3.08 points, SD 7.80, p=0.20.

### IQ tests are not influenced by sleepiness, sleep, or affect

The task scores on verbal, matrix, and block design scores did not show correlations with ESS, CSHQ, actigraphy TST, or PANAS scores in remote or in-person conditions with one exception **(Table 3)**. The CSHQ total score and in-person vocabulary task results showed a trend towards an association (r=-0.28, p=0.06). However, linear regression of in-person vocabulary testing results did not show a main effect for CSHQ total score (F=2.10, p=0.16) when age, gender, and ethnicity were included in the model. All participants reported evening preference so there is not sufficient variability in the data to assess the influence of circadian preference on the cognitive outcomes.

**Table 3:**
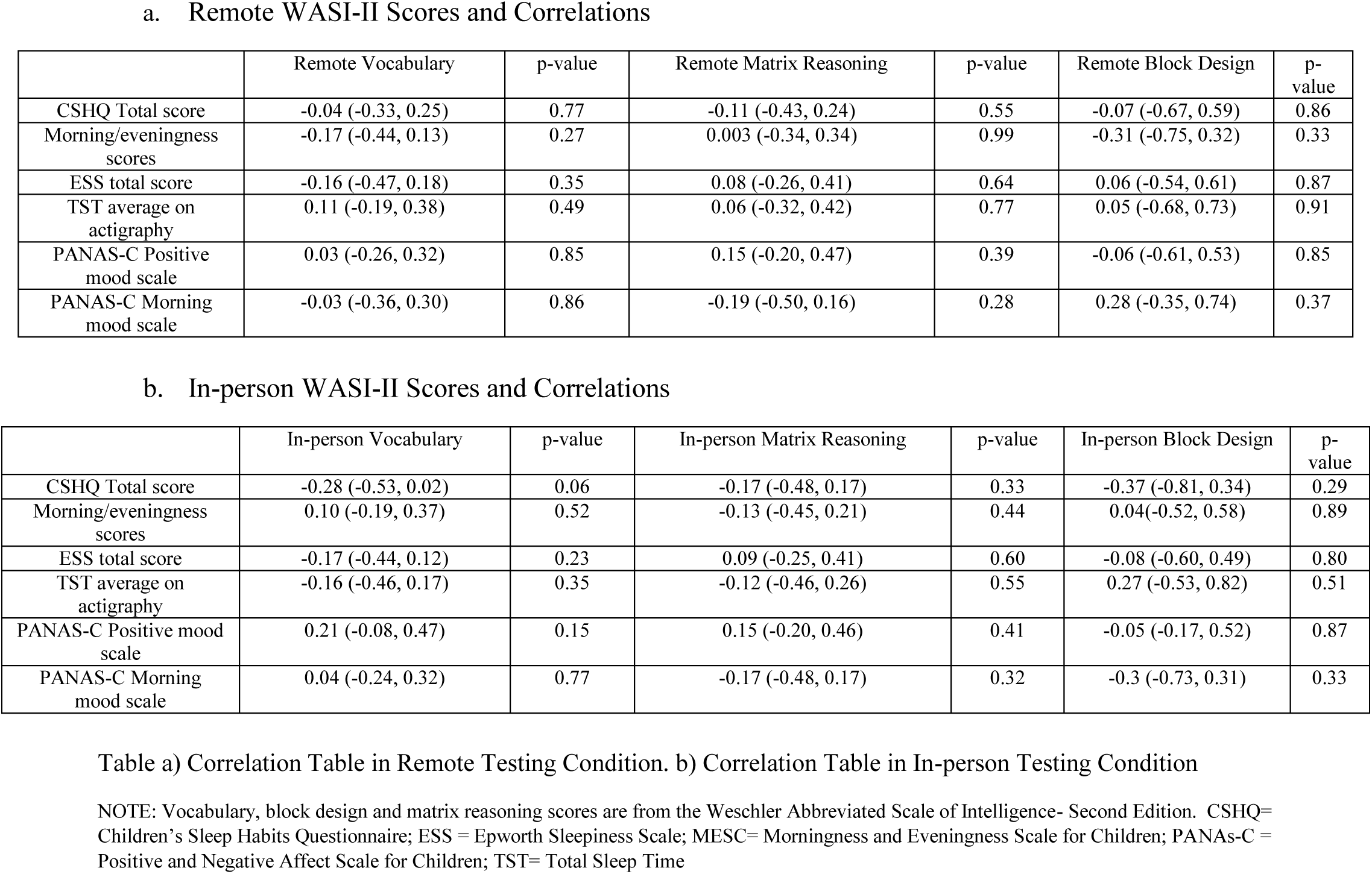
Correlations Tables Between WASI-II Tasks and Sleepiness, Sleep, Affect.

**a. Remote WASI-II Scores and Correlations**
|  | Remote Vocabulary | p-value | Remote Matrix Reasoning | p-value | Remote Block Design | p-value |
| --- | --- | --- | --- | --- | --- | --- |
| CSHQ Total score | -0.04 (-0.33, 0.25) | 0.77 | -0.11 (-0.43, 0.24) | 0.55 | -0.07 (-0.67, 0.59) | 0.86 |
| Morning/eveningness scores | -0.17 (-0.44, 0.13) | 0.27 | 0.003 (-0.34, 0.34) | 0.99 | -0.31 (-0.75, 0.32) | 0.33 |
| ESS total score | -0.16 (-0.47, 0.18) | 0.35 | 0.08 (-0.26, 0.41) | 0.64 | 0.06 (-0.54, 0.61) | 0.87 |
| TST average on actigraphy | 0.11 (-0.19, 0.38) | 0.49 | 0.06 (-0.32, 0.42) | 0.77 | 0.05 (-0.68, 0.73) | 0.91 |
| PANAS-C Positive mood scale | 0.03 (-0.26, 0.32) | 0.85 | 0.15 (-0.20, 0.47) | 0.39 | -0.06 (-0.61, 0.53) | 0.85 |
| PANAS-C Morning mood scale | -0.03 (-0.36, 0.30) | 0.86 | -0.19 (-0.50, 0.16) | 0.28 | 0.28 (-0.35, 0.74) | 0.37 |

**b. In-person WASI-II Scores and Correlations**
|  | In-person Vocabulary | p-value | In-person Matrix Reasoning | p-value | In-person Block Design | p-value |
| --- | --- | --- | --- | --- | --- | --- |
| CSHQ Total score | -0.28 (-0.53, 0.02) | 0.06 | -0.17 (-0.48, 0.17) | 0.33 | -0.37 (-0.81, 0.34) | 0.29 |
| Morning/eveningness scores | 0.10 (-0.19, 0.37) | 0.52 | -0.13 (-0.45, 0.21) | 0.44 | 0.04(-0.52, 0.58) | 0.89 |
| ESS total score | -0.17 (-0.44, 0.12) | 0.23 | 0.09 (-0.25, 0.41) | 0.60 | -0.08 (-0.60, 0.49) | 0.80 |
| TST average on actigraphy | -0.16 (-0.46, 0.17) | 0.35 | -0.12 (-0.46, 0.26) | 0.55 | 0.27 (-0.53, 0.82) | 0.51 |
| PANAS-C Positive mood scale | 0.21 (-0.08, 0.47) | 0.15 | 0.15 (-0.20, 0.46) | 0.41 | -0.05 (-0.17, 0.52) | 0.87 |
| PANAS-C Morning mood scale | 0.04 (-0.24, 0.32) | 0.77 | -0.17 (-0.48, 0.17) | 0.32 | -0.3 (-0.73, 0.31) | 0.33 |

Table a) Correlation Table in Remote Testing Condition. b) Correlation Table in In-person Testing Condition
NOTE: Vocabulary, block design and matrix reasoning scores are from the Weschler Abbreviated Scale of Intelligence- Second Edition. CSHQ= Children's Sleep Habits Questionnaire; ESS = Epworth Sleepiness Scale; MESC= Morningness and Eveningness Scale for Children; PANAs-C = Positive and Negative Affect Scale for Children; TST= Total Sleep Time

## Discussion

Our pilot study results show that use of telehealth to collect verbal and non-verbal IQ scores with may offer a feasible and reliable means to acquire cognitive data for pediatric sleep research through the COVID-19 pandemic and beyond. Between remote and in-person conditions, participants showed good reliability on the vocabulary testing, moderate reliability on the matrix reasoning testing, and poor reliability on the block-design testing. Notably, it was the subjectively sleepy and NT1 participants who showed good to excellent reliability on the vocabulary task whereas healthy controls demonstrated moderate reliability. On matrix reasoning, the subjectively sleepy group again showed excellent reliability on testing conditions; NT1 group and healthy controls showed moderate reliability. Test results did not significantly differ between conditions based on paired t-tests. Last, we did not find that sleepiness, affect, or sleep influenced WASI-II T-scores in either the home or in-person conditions showing IQ is robust to these external factors.

Our results are consistent with other pediatric studies showing high correlation and reliability between remote and in-person verbal and non-verbal cognitive measures. Hodge et al. (2019) assessed 33 children (ages 8 – 12) with learning disabilities using the Wechsler Intelligence Scale for Children-Fifth Edition remotely and in-person and showed very high correlations (r=0.98-0.997) between testing methodologies. Stain et al. (2011) used a broad cognitive testing battery to study remote vs. in-person testing methods among n=11 young people (14-30 years) experiencing psychosis (tests included the Wechsler Test of Adult Reading, WMS-R Logical Memory Subtest, WAI-III Digit Span Subtest, and Controlled Oral Word Association Test).

With the exception of the Digit span task, correlations reported by Stain et al. were significant and r-values ranged between 0.81-0.96. Plausibly, these correlations were higher in these studies than ours because remote testing took place in a testing/clinic site hubs rather than home as in our study. The home environment has more variable quality computers, webcam, and testing environment than hospital/research site testing centers. We did not collect data on participant home network bandwidth speed or their perceived visual and sound quality. Our study adds to the literature in presenting data using interclass correlation testing which takes into account rater bias and is a better assessment of reliability than Pearson or Spearman correlation testing (Shrout & Fleiss, 1979). Another strength of our study is that we report data collected from healthy controls and not just from patient populations. This yielded some surprising findings with the healthy controls showing less reliability in testing methods than patient populations. Though demographic data did not differ between groups, there could be sample bias or unknown confounding factors contributing to these group discrepancies.

While most participant difference scores between remote and in-person assessments were within two standard deviations of the mean on Bland-Altman plots, there were outliers with a maximum difference in measures of 17 on the vocabulary task and 22 on the matrix reasoning task (Figure 1). Such differences would produce changes in the interpretation of results (out of normal range) and thus we suggest investigators continue to refine our testing protocols. WASI-II T-score differences were not significantly different between testing methodologies but vocabulary T-scores were higher in the remote condition than in-person (mean difference 1.08). Similar studies conducted with adult participants have also noted better cognitive test scores using telehealth than in-person and authors suggested that participants may find it easier to focus and feel less pressure to perform when the examiner is not in the room directly observing them (Jacobsen et al., 2003; Kirkwood et al., 2000). However, better remote performance in our study could also be because remote testing tended to occur earlier in the afternoon/evening than in-person assessments and NT1 participants were allowed to stay on wake promoting medications for the remote condition. More standardized timings and conditions of assessments as well as collection of additional measures including momentary fatigue and sleepiness prior to testing in different conditions are recommend for future protocols. In contrast, our participants had higher scores on non-verbal tasks when administered in-person. Methods employed by Hodge et al. (2019) which included split screen mechanisms (one showing the evaluator and other presenting design to replicate) and touch screen technology making it easier to record responses may yield more reliable results on non-verbal tasks (Hodge et al., 2019).

There are additional limitations to our pilot study beyond small sample size. First, we only tested a subset of tests from the WASI-II. Our results may not be applicable when using other cognitive assessments. Second, although all our research assistants were trained to administer the tests in the same way it is possible that there were unstudied differences in administration over the course of the study. Another limitation is the decision to administer the same test within a 1week time span. It is possible that the participants remembered some of the questions; however, our study was counterbalanced to counter learning effects.

## Conclusion

Remote cognitive assessments in patients with sleep disorders and/or daytime sleepiness holds great promise for cognitive sleep research. Importantly, we did not find any associations between WASI-II T-score and subjective or objective nocturnal sleep measures, affect, or baseline sleepiness providing evidence that IQ assessment is robust in both testing conditions. We suggest improvements in our protocols including improved technology (use of split screen and touch screen technology) to assess non-verbal IQ and more standardized assessment protocols overall. While the chosen WASI-II tasks allowed us to obtain baseline IQ scores, they do not give a detailed profile of cognition and functional problems. Reliability of remote testing of cognitive domains such as executive functioning, memory, and attention still need further study in patients with sleep disorders. We hope others build on our study experience as telehealth neuropsychological testing is a necessary need during the COVID-19 pandemic and offers great promise in the future to collect data in less costly and burdensome fashion.

## Data Availability

Data are not accessible at this time

## Notes

Funding Details: This work was supported by grants awarded to Dr. Kiran Maski: Office of Faculty Development Grant from Boston Children’s Hospital and NINDS K23 NS104267-01A1

### Competing Interest Statement

The authors have declared no competing interest.

### Funding Statement

This work was supported by grants awarded to Dr. Kiran Maski from the Office of Faculty Development Grant from Boston Childrens Hospital and NINDS K23 NS104267-01A1

### Author Declarations

IRB approval was obtained from the Boston Childrens Hospital

